# Development and Preliminary Clinical Feasibility of a Wearable Nanovibration Delivery Device for Localised Bone Stimulation in Individuals with Spinal Cord Injury

**DOI:** 10.64898/2026.07.09.26357644

**Authors:** Jonathan A. Williams, Richard Gibson, Paul Campsie, Matthew J. Dalby, John S. Riddell, Mariel Purcell, Sylvie Coupaud, Peter G. Childs, Stuart Reid

## Abstract

Spinal cord injury (SCI) causes rapid and severe bone loss in the paralysed lower limbs, particularly at the distal femur and proximal tibia, where fragility fracture risk is high. *In vitro* nanoscale vibration at 1 kHz has been shown to promote osteogenic differentiation and inhibit osteoclastogenesis, suggesting potential as a targeted mechanical intervention. This study aimed to develop and evaluate a wearable device for delivering and monitoring localised nanovibration at the distal femur in individuals with SCI.

The device delivered continuous sinusoidal nanoscale stimulation at 1 kHz via a bone-conduction transducer, with an opposing accelerometer used to monitor transmitted vibration in real time. Design and target-site selection were refined through two healthy-volunteer investigations comparing the distal femur, proximal tibia, and distal tibia. Bovine femur experiments characterised vibration transmission under controlled benchtop conditions. Preliminary repeated-use feasibility was assessed in one individual with motor-complete SCI.

Healthy volunteer testing showed that although the ankle initially produced the highest transmitted amplitudes, these were highly variable, and positioning was inconsistent. Within the knee region, the distal femur provided the most practical and repeatable site for a wearable application. In bovine femur experiments, scanning laser vibrometry demonstrated measurable vibration on the condylar surface opposite the transducer, and depth-resolved measurements confirmed that nanoscale vibration remained detectable within bone. A gel interface layer reduced the transmitted amplitude. In the feasibility evaluation, 61 sessions were completed over 14 weeks, with logged accelerometry confirming repeated nanoscale vibration transmission.

These findings establish feasibility and support further device optimisation and translational studies.

## 1. Introduction

Spinal cord injury (SCI) is associated with rapid and severe osteoporosis in the paralysed lower limbs, particularly during the early months after injury (Abdelrahman et al., 2021). Bone loss is especially marked at the trabecular-rich ends of the sublesional long bones around the knee and ankle, where fragility fracture risk is markedly increased during routine activities such as wheelchair transfers (Eser et al., 2004; Coupaud et al., 2015; Abdelrahman et al., 2023). These fractures are associated with substantial morbidity and are difficult to manage in the SCI population (Frotzler et al., 2015).

Current strategies for preventing or treating osteoporosis include anti-resorptive and anabolic pharmacological agents, but these approaches have important limitations and do not fully address the clinical problem of fracture-prone bone loss after SCI (Seeman & Martin, 2019; Langdahl, 2021). Mechanical interventions are conceptually attractive because bone is mechanoresponsive (Wang et al., 2022), but many available approaches rely on weight-bearing, active exercise, or whole-limb loading, which may be difficult or impractical in recent motor-complete SCI (Sutor et al., 2022). These limitations highlight the need for alternative non-invasive interventions that can target vulnerable skeletal regions directly.

Nanoscale-amplitude vibration (nanovibration) has shown osteogenic potential in previous *in vitro* studies, where continuous nanovibrational stimulation (1 kHz, 30- or 90 nm amplitude) promoted osteogenic differentiation of human mesenchymal stem cells and supported mineralisation in 3D culture systems (Nikukar et al., 2013; Tsimbouri et al., 2017; Orapiriyakul et al., 2020). Further, nanovibrational stimulation of co-cultures of osteoblast (bone-forming) and osteoclast (bone-resorbing) progenitor cells simultaneously enhanced osteogenesis while inhibiting the formation of new osteoclasts (Kennedy et al., 2021). More recently, nanovibrational stimulation has also been reported to enhance osteogenic responses *in vivo* in zebrafish, increasing osteoblast number and bone formation during larval development and increasing osteoblast reporter activity at adult scale injury sites, while producing comparatively limited effects in non-skeletal tissues (Adigun et al., 2026). These findings suggest that localised nanovibration may offer a mechanically targeted approach for stimulating bone-relevant responses. However, translating this concept from a cell culture system (Campsie et al., 2019), for direct *in vivo* or clinical use, is not straightforward and would require a wearable system capable of delivering controlled nanovibration to a fracture-prone skeletal site while simultaneously monitoring the transmitted signal in real time, to ensure vibration delivery.

In this respect, the present approach differs from platform-based interventions, such as low-magnitude mechanical signals (also referred to as low-intensity vibration) and whole-body vibration, which expose the body or lower limbs to dynamic mechanical loading and may involve both direct skeletal vibration and muscle-mediated loading through postural and reflex responses (Rubin et al., 2004; Cardinale & Rittweger, 2006; Divasta et al., 2024). It also differs from functional electrical stimulation approaches that have been applied in SCI, which act more explicitly by electrically induced muscle contractions, thereby generating muscle- and tendon-mediated skeletal loading (Shields & Dudley-Javoroski, 2007). Pressure-wave-based interventions, such as low-intensity pulsed ultrasound and shockwave therapy, provide further examples of mechanically based bone stimulation, but are generally delivered using external clinical applicators rather than wearable devices (Mittermayr et al., 2021; Padilla et al., 2014). By contrast, the present device is not intended to activate muscle, induce joint motion, or generate whole-limb loading, but was instead motivated by the prior *in vitro* nanovibration studies and designed to deliver a localised, wearable, continuous sinusoidal 1 kHz nanoscale stimulus directly to the target skeletal region.

In previous work, we developed a wearable nanovibration delivery device for a rat model of complete SCI and showed that externally applied 1 kHz nanovibration could be transmitted into the paralysed hindlimb long bones (Williams et al., 2024). Although the intervention did not reverse the severe established osteoporosis in that model (Williams et al., 2020, 2022, 2024), it demonstrated the feasibility of targeted bone-level nanovibration delivery *in vivo* and was associated with an increase in the bone formation marker P1NP, suggesting stimulation of early bone-forming activity.

For translation to human use after SCI, successful wearable delivery depends not only on the biological rationale for nanovibration, but also on solving key engineering problems, including anatomical target-site selection, repeatable positioning, interface conformity, safe prolonged wear, portability, and confirmation that measurable vibration can be transmitted to and through the target region. Candidate lower-limb sites around the knee and ankle are clinically relevant and externally accessible, but may differ substantially in fit, stability, and measurable vibration transmission when implemented in a wearable format.

The aim of this study was therefore to develop and evaluate a wearable nanovibration device for localised application to the lower limb in participants following SCI. Specifically, we sought to (i) refine device design and target-site selection through testing in healthy volunteers, (ii) characterise transmission of externally applied nanovibration through bovine femur bone under controlled benchtop conditions, and (iii) assess the preliminary clinical feasibility of repeated device use in individuals with motor-complete SCI.

## 2. Methods

### 2.1 Device design and operating principle

A wearable nanovibration device was developed to deliver controlled, sinusoidal, nanoscale-amplitude mechanical stimulation at 1 kHz to clinically relevant lower-limb skeletal sites and to monitor transmitted vibration during use. It was intended for use in individuals with motor-complete traumatic spinal cord injury (SCI) at neurological level C4 or below, recruited as soon as feasible after injury. These individuals have a complete loss of motor function and sensation below the neurological level of injury, and are wheelchair users and therefore vulnerable to pressure-related skin injury at the device interface. The principal design requirements were therefore that the device should (i) deliver a consistent vibration to a target bone region, (ii) measure and record the vibration transmitted to the opposite side of that region, and (iii) permit repeated use without causing local tissue damage. To reduce this risk, compliant padding was incorporated into the contact surfaces of both the transducer, accelerometer and strap arms (Figure 1A). The device architecture was designed to support evaluation of candidate lower-limb target sites, with component placement guided by accessible external bony landmarks.

**Figure 1.**
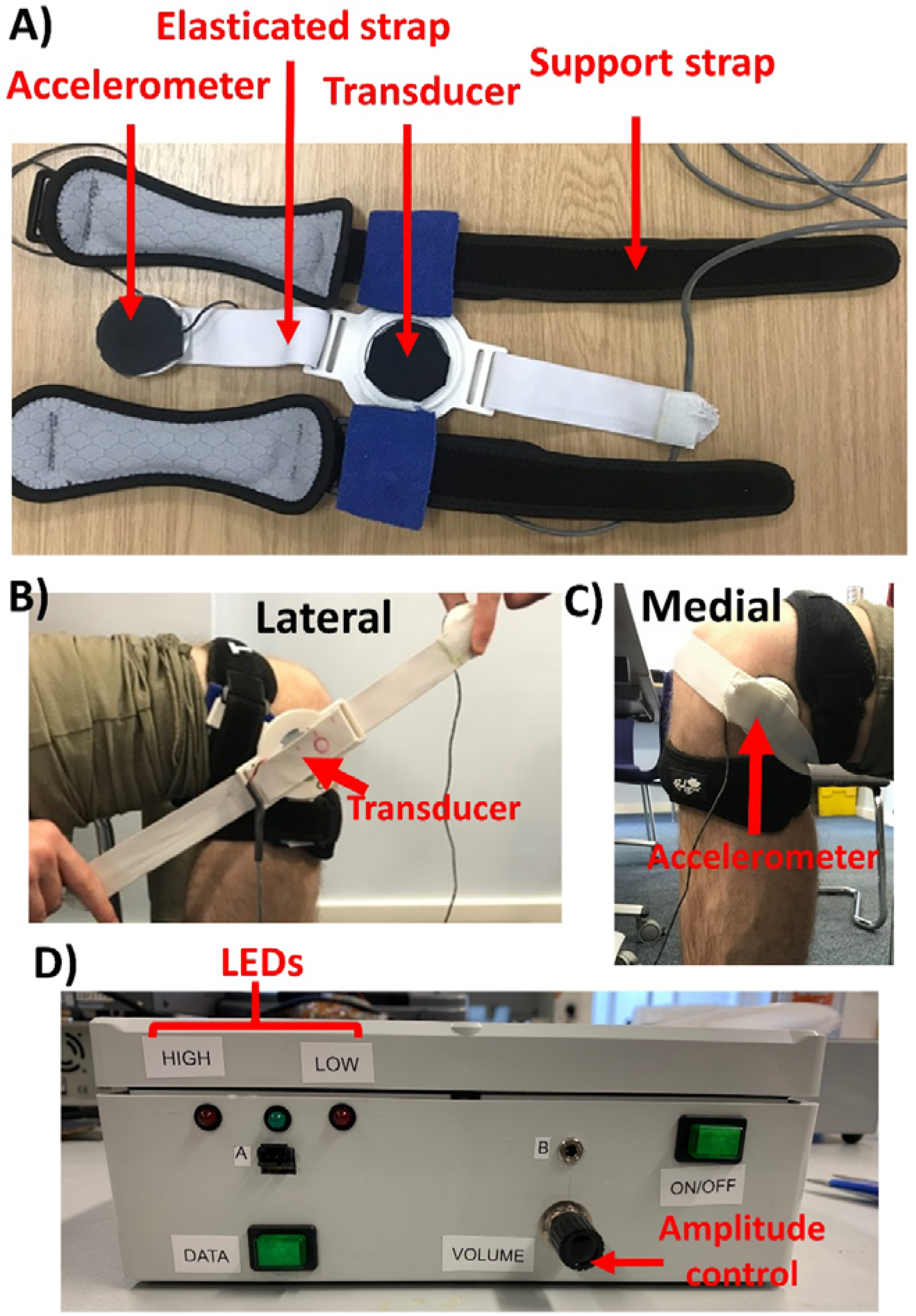
Design and final configuration of the wearable nanovibration delivery device. (A) Final assembled wearable device prior to application, including the central device hub, modular arms, support straps, elasticated strap, transducer, accelerometer, and gel interface layer. (B) Device application with the knee in a flexed position, showing the modular-arm configuration used to secure the transducer laterally across the distal femur. (C) Medial view of the device in position, showing the accelerometer located opposite the transducer for measurement of vibration transmitted across the target region. (D) A portable electronics unit used to generate the 1 kHz drive signal, control output amplitude, provide LED feedback on transmitted vibration level, and log accelerometer data during use.

The wearable system comprised a bone-conduction transducer, a piezoelectric accelerometer, bespoke 3D-printed housings, modular support arms, fixation straps, and a custom electronics unit (Figure 1A). In its final configuration for clinical use, the vibration source was a large-surface LB07 bone conduction transducer with a 44 mm diameter contact surface, selected to apply vibration over a relatively broad skin-contact area, improving coupling to the underlying tissues. The transducer was positioned on the lateral aspect of the distal femur (Figure 1B), while an ACH-01 piezoelectric polymer film accelerometer was positioned on the medial side opposite the transducer to detect the vibration transmitted across the target region (Figure 1C). This lateral–medial arrangement allowed the target bone region to lie between the source and sensing components, enabling real-time monitoring of transmitted vibration during use. Both components were mounted in custom 3D-printed housings connected to modular arms and secured to the limb using a combination of non-elastic support straps and an elasticated strap to maintain alignment, fixation, and contact pressure. A 9 mm-thick layer of a proprietary polyurethane gel interface material (Technogel®, Technogel Germany GmbH), commonly used for pressure-distributing padding in orthotic and prosthetic applications, was incorporated at the skin–device interface (both rigid transducer and accelerometer surfaces) to reduce local pressure and improve conformity during wear.

The electronics system was designed to generate a controlled 1 kHz sinusoidal drive signal for the bone conduction transducer and to process and log the accelerometer output in a portable patient-use configuration (Figure 1E). Wave generation was based on an AD9833 programmable waveform generator controlled by an ATMega328 microcontroller. The output signal was filtered with a seventh-order LC elliptical reconstruction filter and amplified with OPA37-based amplifier stages, with user-adjustable output control via a rotary potentiometer before final power amplification to drive the transducer. The ACH-01 accelerometer signal was processed using a low-noise multistage amplifier and converted to a DC signal using an AD736JNZ RMS-to-DC converter before being read by an Arduino Nano. The Arduino applied an accelerometer-specific calibration equation to convert the measured voltage into displacement amplitude, logged these data to a micro-SD card, and controlled three LED indicators that provided real-time user feedback on whether the transmitted vibration amplitude was within the desired operating range. One green LED indicated in-range vibration amplitude, and two red LEDs indicated that the transmitted amplitude was below or above the target range (< 1.5 nm or > 40 nm) (Figure 1D). The system was powered by a rechargeable 5 V portable battery pack, enabling stand-alone operation without mains power during the intervention. The electronics were housed in a portable external unit (Figure 1D) that could be positioned beneath the wheelchair in the space below the participant’s legs, supporting device operation during wheelchair use and outside a fixed bedside setting. A more detailed overview of the electronics system, including a description of accelerometer calibration, is available in Supplementary Material A.

In operation, the transducer delivered vibration from the lateral side of the joint, and the accelerometer measured the transmitted vibration on the medial side, allowing the device position or output amplitude to be adjusted in response to real-time feedback.

### 2.2 Device development and iterative testing in healthy volunteers

The wearable device design was refined through two iterative investigations in healthy adult volunteers at the University of Strathclyde. These studies were undertaken to (i) quantify the amplitude and consistency of vibration transmission at candidate lower-limb target sites, (ii) obtain user and operator feedback to improve the device design, and (iii) select the most appropriate anatomical target region for the preliminary clinical feasibility evaluation. Because early SCI-induced osteoporosis affects fracture-prone sublesional sites in the lower limb, candidate target sites were selected around the knee and ankle regions. The target sites evaluated were the distal femur, proximal tibia, and distal tibia. For each site, the transducer and accelerometer were positioned using palpable external bony landmarks, namely the distal femoral condyles, proximal tibial condyles, and distal tibial malleoli. The studies were not intended to assess any biological effect of nanovibration, but rather to compare device fit and measurable transmission at these regions across different individuals.

Both investigations were approved by the Departmental Ethics Committee at the University of Strathclyde. Healthy adult volunteers were recruited from the University of Strathclyde. Participants were excluded if they were younger than 18 years, had a musculoskeletal condition or disability affecting the lower limb, inflammatory joint disease, an implant in the knee or ankle region, an inflammatory skin condition, pregnancy, or recent lower-limb surgery.

In the initial investigation, the prototype device was applied sequentially to the distal femur, proximal tibia, and distal tibia. During testing, participants were seated with the right knee and ankle exposed, and the investigator identified the most appropriate local application position by palpation of the relevant bony landmarks. The transducer was placed on the lateral side of the selected site, and the accelerometer on the opposite side. The early prototype was powered using a 20 V peak-to-peak sinusoidal signal at 1 kHz. Nanovibration was delivered for no more than 5 minutes at each site, and the transmitted vibration amplitude was recorded using the accelerometer. Participants were asked to remain still and minimise lower-limb movement during measurement.

A follow-up investigation was then performed using a revised prototype incorporating design changes informed by the first phase. These modifications included a dedicated accelerometer housing, revised strap routing through the device arms, rounded housing edges, and a redesigned ankle harness intended to improve repeatable positioning at the malleolus. The same anatomical sites, general setup, and measurement approach were used as in the initial investigation. Representative device configurations are shown in Supplementary Material B.

The distal tibia was ultimately not taken forward to the clinical feasibility study due to practical difficulties in achieving stable, repeatable contact at the malleolus across different individuals. These difficulties were attributed to the anatomical variability and geometric complexity of the ankle region, which limited both consistent fit and reproducible vibration delivery. Within the knee region, the healthy volunteer investigations indicated that the distal femur provided the most suitable balance of accessibility, fit, and measurable vibration transmission for subsequent clinical use. In addition, the geometry of the flexed knee made this site more practical for external device attachment than the proximal tibia, allowing more stable positioning and strap fixation.

### 2.3 Bovine femur transmission experiments

Bovine femur experiments were performed to characterise the transmission of externally applied nanovibration through bone under controlled laboratory conditions. Juvenile bovine femora were selected as a surrogate for human bone because they are readily available, represent large weight-bearing long bones of relevant scale, and are commonly used in biomechanical research as a practical proxy for human femoral tissue (Fletcher et al., 2018).

Femora were sourced from the John Scott Abattoir (Paisley, Scotland) and stored at −20 °C. Samples were limited to a single freeze-thaw cycle. For the main transmission experiments, only the knee-joint end of each femur was retained. Excess soft tissue was removed manually, and the proximal portion of the bone was sawn away while retaining at least 5 cm of shaft to allow secure clamping and attachment of the vibration device (Figure 3A).

A small set of preliminary scanning laser interferometry experiments was first performed to visualise the spread of vibration across the bovine condylar surface (Figure 3A). The bone was clamped in front of a scanning laser interferometer (PSV-500-H, Polytec, Germany), and measurements were acquired on the exposed condylar surface directly opposite the transducer. The interferometer bandwidth was set above 2.5 kHz to capture the 1 kHz input signal, and these experiments were repeated on two bovine samples.

For the main depth-resolved transmission experiment, seven bovine femora were prepared. The wearable nanovibration device used in the clinical study was adapted for benchtop use by removing the accelerometer housing, modular arms, and standard support straps, after which the device was secured to the distal femoral condyle using an elasticated strap adapted to the geometry of the bovine sample. The same voice-coil bone-conduction transducer used in the wearable device was driven by a 20 V peak-to-peak, 1 kHz signal from a signal generator. A single-point laser interferometer (SIOS Meßtechnik GmbH, Germany) was used for the main transmission measurements, and retroreflective tape was applied to the bone surface and later within drilled measurement sites to improve signal quality. The experimental setup is shown in Supplementary Material C.

For depth-resolved measurements, the femur was clamped with the condylar region facing the laser interferometer. Surface measurements were first taken on the far side of the condyle opposite the transducer. A 1 cm diameter flat drill bit was then used to drill into the condylar region, allowing vibration measurements to be taken at 1 cm depth increments through the drilled region. At each increment, retroreflective tape was inserted into the drilled hole and vibration amplitude at 1 kHz was measured using the interferometer. Measurements were repeated at successive depths until the bone was fully traversed. A further set of measurements was taken from the transducer surface once the drilled channel had fully penetrated the bone, to allow laser access while the device remained coupled to the bone. These measurements were used to normalise internal bone vibration amplitudes to the transducer output. Each set of measurements, including bone surface, internal, and transducer-surface measurements, was repeated five times and averaged. At each stage, measurements were made with and without the gel interface layer used in the preliminary clinical feasibility evaluation.

Further, to assess repeatability of surface-level transmission following repeated device placement, an additional repeat-application experiment was performed on a single bovine femur sample. The device was applied and removed repeatedly six times, with and without the gel interface layer. For each application, the transmitted vibration amplitude at 1 kHz was measured five times on the bone surface opposite the transducer using the single-point laser interferometer. This experiment was intended to capture variability associated with repeated reapplication of the device, including small differences in positioning, local conformity, strap tension, and interface behaviour.

### 2.4 Preliminary clinical feasibility evaluation in SCI

A preliminary clinical feasibility evaluation was conducted at the Queen Elizabeth National Spinal Injuries Unit, Glasgow, UK, to assess whether the wearable nanovibration device could be applied and used repeatedly by individuals with motor-complete spinal cord injury while maintaining measurable transmitted vibration at the target site. Ethical approval was obtained from NHS Greater Glasgow and Clyde / West of Scotland REC 3 (REC reference 15/WS/0009; IRAS project ID 84046), including approval of an amendment to introduce localised vibration using bone-conduction transducers. All participants provided informed consent prior to participation.

Participants were recruited from inpatients at the Queen Elizabeth National Spinal Injuries Unit. For the intervention study, eligible participants had motor-complete traumatic SCI at neurological level C4 or below and were recruited as soon as feasible after injury. General exclusion criteria included age below 16 years, ventilator dependence, pregnancy, extensive spasticity, bilateral metal implants and/or recent fractures involving the tibia, fibula, or femur, and prior medical treatment known to affect bone metabolism. Additional exclusion criteria for the vibration intervention included acute thrombosis, acute lower-limb tendinopathy, rheumatoid arthritis, epilepsy, pressure sores, psychotic illness or neurotic disturbance, and recent autonomic dysreflexia.

The device was positioned on the right leg, with the transducer placed on the lateral aspect of the distal femur and the accelerometer positioned medially. Participants entering the vibration intervention were instructed in the safe application and operation of the device and were provided with a user manual (Supplementary Material D). The intervention was designed to be delivered while the participant remained seated in their wheelchair. Session duration was increased progressively, beginning at 15 minutes and increasing over subsequent sessions toward a target of up to 4 hours per session, with a planned frequency of up to 5 sessions per week during the intervention phase. In the wearable-device feasibility evaluation reported here, transmitted vibration was monitored during each session, and participants were instructed to use the real-time LED feedback indicators to maintain the signal within the desired operating range by adjusting device position, strap tension, or output amplitude, as required.

The accelerometer output was logged during each session using the on-board data acquisition system. In addition, participants kept a written record of session duration, dates of use, and any practical problems encountered during wear, including interrupted or abandoned sessions. Skin was checked regularly during intervention sessions for evidence of pressure marks at the application sites, in line with the study protocol, and the intervention was to be paused or stopped if persistent pressure marks occurred. The primary purpose of this evaluation was to assess repeated usability, maintenance of transmitted vibration at the target site, and practical feasibility in the SCI setting, rather than to determine clinical efficacy.

Peripheral quantitative computed tomography scans (XCT 3000, Stratec Medizintechnik GmbH, Germany) were planned within 6 weeks of injury and again at 4, 8, and 12 months post-injury as part of the wider longitudinal study. However, recruitment to the wearable-device intervention was limited, and only one participant completed the full evaluation due to disruptions caused by the COVID-19 pandemic; consequently, imaging outcomes are not reported.

### 2.5 Outcome measures

Outcome measures were defined according to the three components of the study: healthy volunteer device development, bovine femur transmission experiments, and preliminary clinical feasibility evaluation in SCI.

For the healthy volunteer investigations, the primary outcomes were the measured amplitude of transmitted vibration at 1 kHz at each candidate anatomical site and the consistency of transmission across repeated applications and participants. These data were used to compare the distal femur, proximal tibia, and distal tibia as candidate sites for wearable device application, and to inform iterative design refinement.

For the bovine femur experiments, the primary outcomes were vibration amplitude at 1 kHz measured on the bone surface and interior at progressive depth intervals and on the transducer surface during active vibration delivery. These measurements were used to assess the transmission of transmitted vibration through bone. Additional comparisons included the effects of the gel interface padding layer and repeated device reapplication on transmitted vibration.

For the preliminary clinical feasibility evaluation, the primary outcomes were session completion, session duration, logged transmitted vibration amplitude at 1 kHz, total intervention time and practical usability of the device during repeated use. Feasibility was assessed in terms of whether measurable transmitted vibration could be maintained at the target site across sessions, whether the device could be applied and used repeatedly in the SCI setting, and whether any practical or skin-related problems limited use.

### 2.6 Statistical analysis

All statistical analyses were performed in Python (v3.10) using standard scientific computing libraries. A *p*-value < 0.05 was considered statistically significant.

For healthy volunteer testing, a repeated-measures framework was used, with vibration amplitude measured at multiple anatomical sites within the same participants. Amplitudes were summarised as mean ± standard deviation (SD) and additionally visualised using median, interquartile range, and individual participant data points. Overall differences between anatomical sites were assessed using repeated-measures ANOVA, with paired *t*-tests and Holm correction used for post hoc comparisons where appropriate. Because some datasets, particularly ankle measurements in the initial investigation, showed substantial variability and skew, Friedman tests were also performed as non-parametric sensitivity analyses. Normality of paired differences was assessed using the Shapiro–Wilk test.

For the depth-resolved bovine femur experiment, repeated interferometric measurements at each depth were first averaged within each bone. Data for the with-gel and without-gel interface conditions were summarised across bones as mean ± SD and plotted as a function of depth. Mean transmitted amplitude across depth was compared between conditions using paired *t*-tests. Depth dependence was assessed by linear regression within each bone, with slope values tested against zero using one-sample *t*-tests and compared between conditions using paired *t*-tests. For the repeat-application experiment, a single bovine femur sample was tested repeatedly under both interface conditions. Mean transmitted amplitude per application and within-application SD were compared between conditions using paired *t*-tests. Coefficients of variation were calculated descriptively, and between-application variability was summarised as the SD of the application means within each condition.

Clinical feasibility data from the wearable device evaluation were analysed descriptively. Session number, session duration, calendar timing of sessions, and logged transmitted vibration amplitude were summarised using means, SDs, and standard errors where appropriate, and visualised to illustrate session adherence and the stability of transmitted vibration over repeated use and across session time. No formal inferential statistical testing was performed for these data because only one participant completed the evaluation.

## 3. Results

### 3.1 Device development and iterative testing in healthy volunteers

Two iterative healthy-volunteer investigations were performed at 1 kHz to compare measurable vibration transmission at candidate lower-limb target sites and to refine the wearable device design.

In the initial investigation, transmitted vibration amplitude was measured at the distal femur, proximal tibia, and distal tibia in 12 participants. Mean transmitted amplitudes at the distal femur and proximal tibia were similar, at 3.0 ± 1.1 nm and 3.2 ± 1.7 nm, respectively, whereas the distal tibia showed a substantially higher mean amplitude of 14.5 ± 13.4 nm (Figure 2A). However, distal tibia measurements were also much more variable than those recorded at the knee sites. Repeated-measures ANOVA demonstrated a significant effect of anatomical site (*p* = 0.0020), with post hoc testing showing no significant difference between the distal femur and proximal tibia, but significant differences between the distal tibia and both. A Friedman test supported the same conclusion (*p* = 0.00019). Although the ankle produced the highest transmitted amplitudes, device application at this site was less repeatable due to poor conformity between the transducer surface and the malleolar region, as well as practical difficulties with strap application and accelerometer positioning.

**Figure 2.**
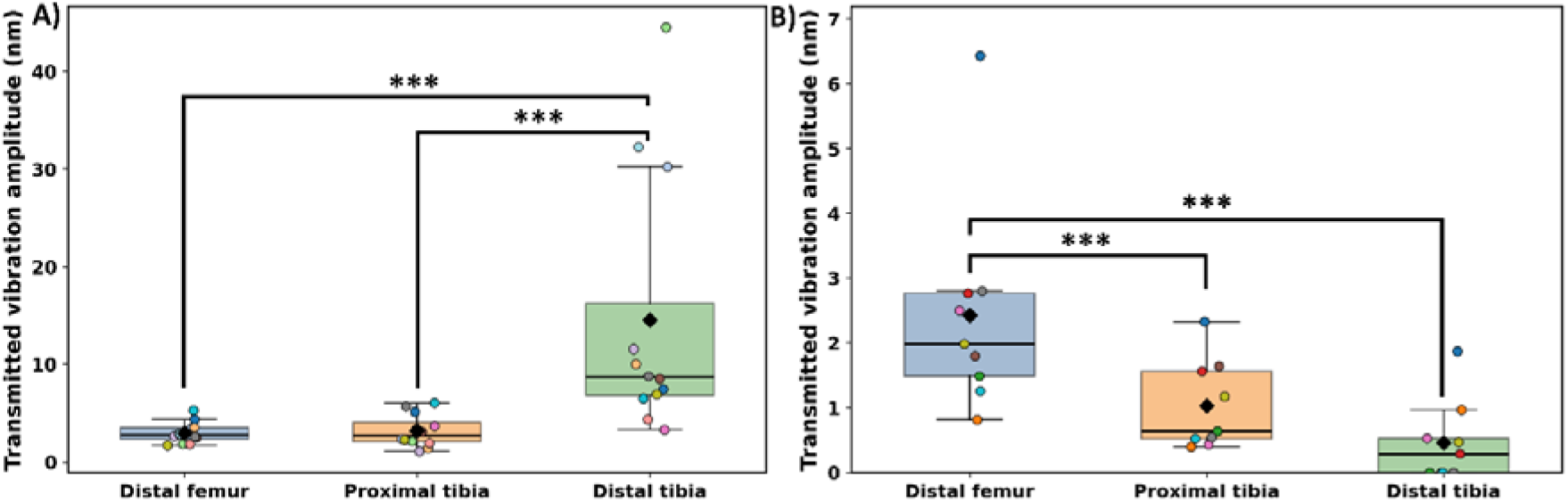
Transmitted vibration amplitude at candidate lower-limb target sites during healthy-volunteer device testing at 1 kHz. **(A)** Initial investigation (12 participants) showed transmitted vibration amplitudes measured at the distal femur and proximal tibia (knee), and distal tibia (ankle). **(B)** Follow-up investigation using the revised prototype (9 participants). In both panels, individual data points are shown together with median, interquartile range, and mean marker (black diamond).

Following the device redesign, a second healthy volunteer study was conducted with 9 participants. The revised prototype incorporated an accelerometer housing, revised strap routing through the device arms, rounded housing edges (Figure 1), and a redesigned ankle harness (Supplementary Material B). In this follow-up investigation, the distal femur showed the highest transmitted amplitude (2.4 ± 1.7 nm), followed by the proximal tibia (1.0 ± 0.7 nm) and distal tibia (0.5 ± 0.6 nm) (Figure 2B). Compared with the initial investigation, overall transmitted vibration amplitude and variability decreased, and the extreme ankle values were no longer observed. Repeated-measures ANOVA again showed a significant effect of anatomical site (*p* = 0.0002), and pairwise comparisons indicated that the distal femur differed significantly from both the proximal tibia and distal tibia. The Friedman test supported the same overall conclusion (*p* = 0.0031). Taken together, these findings indicated that the revised device achieved more consistent vibration transmission and helped identify the distal femur as the most suitable anatomical target site for subsequent preliminary clinical feasibility testing. Further, the distal femur remained easier for consistent device position than the proximal tibia because the geometry of the flexed knee permitted more stable device attachment and strap fixation. The volunteer investigations also informed the final device design, including adoption of a dedicated accelerometer housing, simplified strap routing, and a target operating amplitude of 1.5 nm for subsequent clinical use.

### 3.2 Bovine femur transmission experiments

#### 3.2.1 Surface mapping of transmitted vibration on bovine femur

Preliminary scanning laser interferometry demonstrated that externally applied nanovibration at 1 kHz was measurable on the condylar surface opposite the transducer, confirming transmission across the bovine femur under controlled benchtop conditions (Figure 3A). The mapped response was spatially distributed across the opposing condylar surface, with the highest amplitudes occurring in the region directly opposite the transducer and progressive attenuation away from this region. Peak transmitted surface amplitude was approximately 12 nm, and the mapped signal decreased to approximately half of this value within about 4 cm of the maximum-amplitude region.

#### 3.2.2 Depth-resolved transmission through drilled bovine femur

The wearable device used in the preliminary clinical feasibility evaluation incorporated a compliant gel interface layer to reduce the risk of pressure-related tissue damage. Accordingly, the bovine femur depth-resolved transmission experiment was performed to assess the effect of this interface on vibration transmission at 1 kHz (Figure 3B). Externally applied vibration was measurable at successive 1 cm depth increments through the drilled condylar region, with transmitted amplitudes at 1 kHz remaining within the nanoscale range throughout the region.

**Figure 3.**
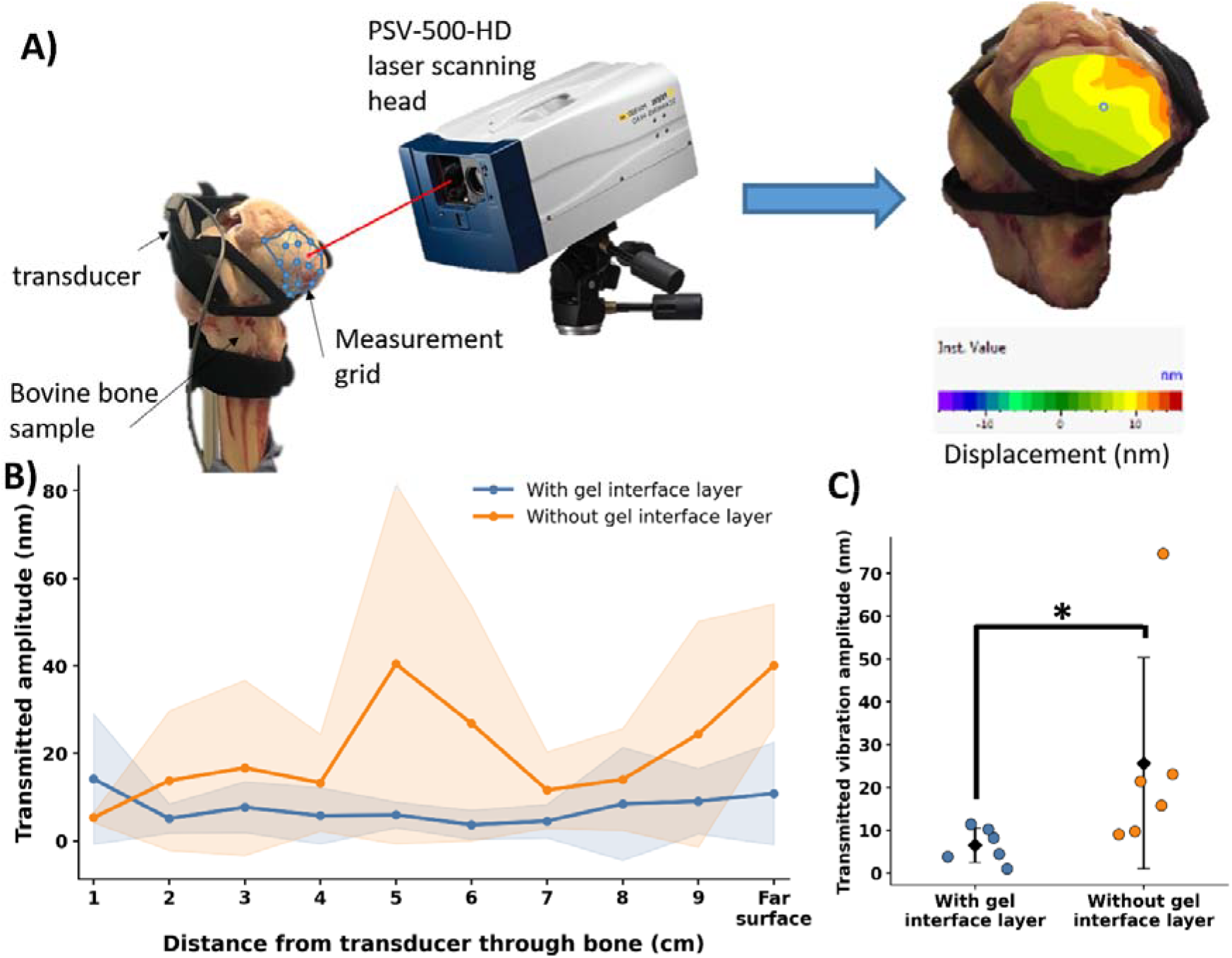
Transmission of 1 kHz nanovibration through bovine femur and effect of the gel interface layer. (A) Experimental arrangement for laser Doppler vibrometry measurements on bovine distal femur samples. A transducer was strapped to the condylar surface, and transmitted vibration was measured using a PSV-500-HD scanning laser vibrometer. A measurement grid was applied over the condylar region to generate displacement maps. A representative displacement map is shown, illustrating spatially resolved nanoscale vibration across the bone surface. (B) Depth-resolved transmission through the drilled bovine femur at 1 kHz, measured at successive 1 cm increments through the condylar region and plotted as distance from the transducer through the bone. Mean transmitted amplitudes across bones are shown for conditions with and without the gel interface layer, with shaded bands indicating standard deviation. (C) Repeat-application experiment on a single bovine femur sample, showing transmitted vibration amplitude measured on the bone surface opposite the transducer following repeated device attachment with and without the gel interface layer. Individual points represent the mean of repeated interferometric measurements for each application, black diamonds indicate condition means, and error bars indicate between-application standard deviation.

The mean transmitted amplitude across depth was significantly lower in the presence of the gel interface (approximately 7 nm) than in the no-gel condition (approximately 21 nm) (paired *t*-test, *p* = 0.0405) (Figure 3B). In the no-gel condition, transmitted amplitude showed a significant positive association with distance from the transducer (one-sided p = 0.0428), whereas no significant distance-dependent association was detected in the gel condition (one-sided p = 0.2466). However, the difference in attenuation slope between conditions was not statistically significant (p = 0.1362). Variability also appeared lower in the gel condition, likely reflecting improved conformity at the transducer–bone interface and more consistent mechanical coupling. These findings show that the presence of the gel interface reduced the overall magnitude of transmitted vibration at 1 kHz.

Transmitted amplitudes at 1 kHz were also normalised to the corresponding transducer surface amplitude measured under each interface condition, while the device was coupled to the bone (Supplementary Material E). In the no-gel condition, on average, 1.3% of the transducer surface amplitude was detected within the bone across the measured depth range, compared with 0.4% in the gel-interface condition.

#### 3.2.3 Device attachment repeatability

To further assess the repeatability of surface-level transmission following repeated device attachment, the device was applied and removed six times on a single bovine femur sample, with and without the gel interface layer (Figure 3C). Transmitted amplitude across repeat attachment was lower with the gel interface layer than without it (paired t-test, one-sided p = 0.0461), consistent with the depth-resolved experiment. Raw within-application standard deviation was significantly lower with gel than without gel (paired t-test, one-sided p = 0.0199), and between-application variability of the application means was also lower with gel (SD = 4 nm) than without gel (SD = 24 nm). However, because the mean transmitted amplitudes differed substantially between conditions, comparisons based on raw SD should be interpreted cautiously. Consistent with this, relative variability, assessed by the coefficient of variation, did not differ significantly between conditions (p = 0.4396). Taken together, these results are consistent with, but do not conclusively demonstrate, more reproducible mechanical coupling when the compliant gel interface layer was present.

### 3.3 Preliminary clinical feasibility evaluation in SCI

#### 3.3.1 Session duration and adherence

Recruitment to the wearable-device intervention was limited, and only one participant completed the full feasibility evaluation. Accordingly, the results are presented descriptively as a repeated-use feasibility dataset rather than as a formal efficacy analysis. The participant completed 61 vibration sessions between November 2019 and February 2020, spanning approximately 14 weeks and totalling 119.4 hours of intervention. The pattern of use was broadly consistent with the intended weekday-based schedule (4.3 sessions per week, on average), although intermittent gaps between sessions were present. Session duration increased during the early familiarisation phase but did not reach the originally planned 4 h maximum, as discussed in Section 3.3.3. Instead, most sessions stabilised at approximately 2 h in duration (Figure 4A,B). The mean session duration across all sessions was 117.4 min.

**Figure 4.**
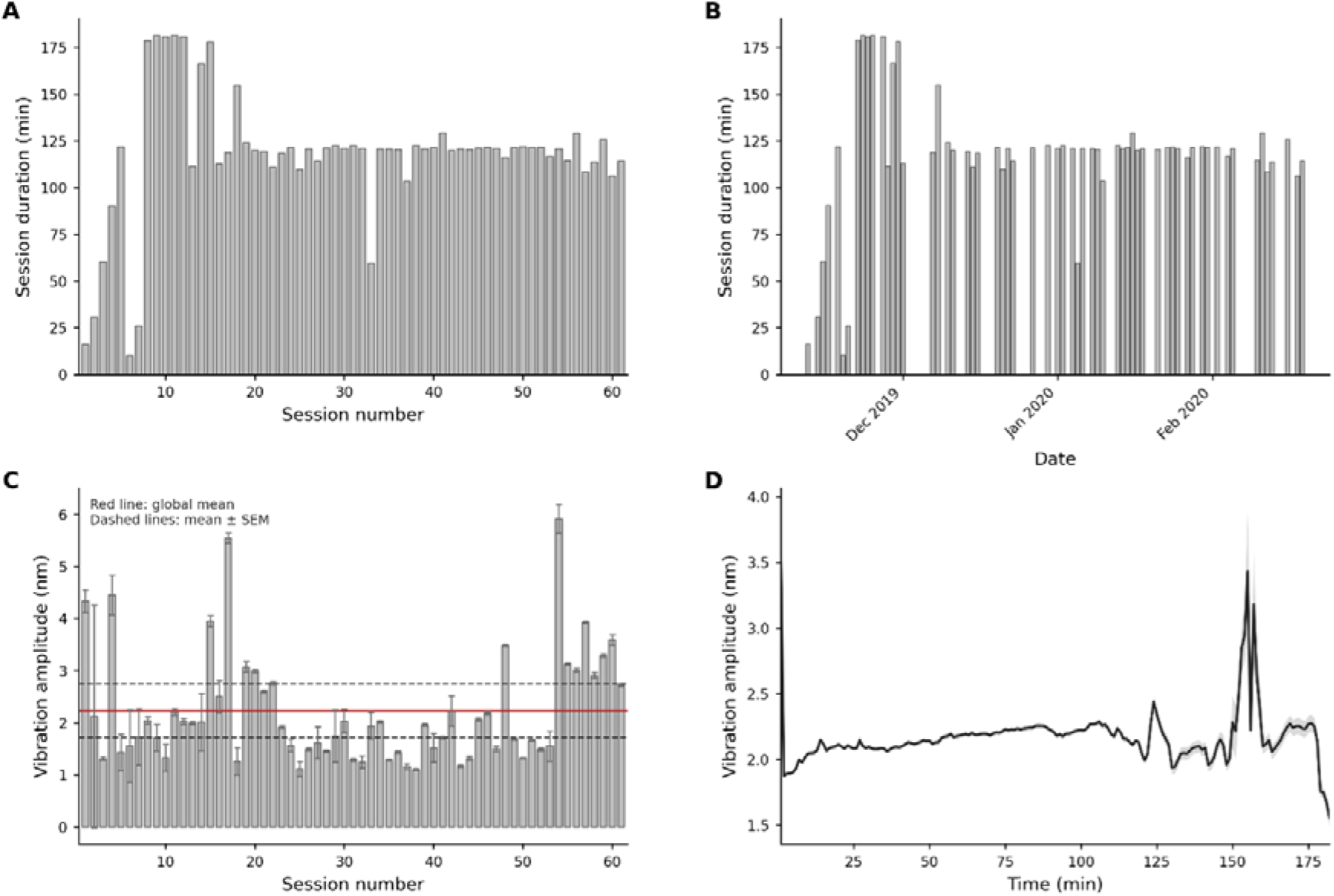
Preliminary clinical feasibility evaluation of the wearable nanovibration device in a participant with motor-complete spinal cord injury. **(A)** Session duration as a function of session number. **(B)** Session duration over calendar time, with gaps representing days without recorded sessions. **(C)** Mean transmitted vibration amplitude at 1 kHz per session with standard deviation error bars; the red line denotes the overall mean and dashed lines denote the corresponding standard error bounds. **(D)** Mean transmitted vibration amplitude at 1 kHz across session time, averaged across sessions, with standard error shading.

#### 3.3.2 Logged transmitted vibration during use

Logged accelerometry confirmed that transmitted vibration was maintained at the distal femur across repeated sessions. The overall mean transmitted vibration amplitude across sessions was 2.23 nm, indicating that recorded transmission was generally maintained above the minimum acceptable threshold of 1.5 nm for device operation (Figure 4C). Although some sessions showed higher average values and greater within-session variability than others, the averaged signal remained broadly stable for most of the session duration (Figure 4D). Transient increases were observed at the beginning and toward the end of sessions and were associated with device attachment and removal, respectively.

#### 3.3.3 Practical usability observations

The participant’s usage log showed that the device could be used repeatedly over an extended period, though not every day, with breaks between sessions (Figure 4B). No pressure sores or persistent skin damage were observed at the device interface sites over the 61 sessions. Practical limitations identified during the feasibility evaluation included gradual device slippage during prolonged wear, audible hum from the transducer (1 kHz is audible), bulk of the external electronics unit and cabling, and difficulty wearing the device discreetly under clothing. These factors reduced convenience and likely contributed to the session being shorter than planned. The device could also be used with the leg extended while lying down by removing one modular arm, and this was achieved successfully without reported difficulty.

## 4. Discussion

This study describes the development of a wearable nanovibration delivery device designed for localised application to the distal femur in patients with complete spinal cord injury (SCI), and evaluates its performance through healthy-volunteer testing, bovine bone transmission experiments, and a preliminary clinical feasibility study. The principal findings were that the distal femur emerged as the most suitable target site for repeatable wearable application, externally applied nanovibration at 1 kHz was measurable across and within bovine femur bone under controlled benchtop conditions, and repeated device use with maintained nanoscale transmitted vibration was feasible in an individual with motor-complete SCI. Together, these findings support the translational feasibility of a wearable system for delivering and monitoring localised nanovibration at a fracture-prone skeletal site, while also highlighting important practical and engineering constraints that must be addressed before larger clinical studies can be justified.

The healthy-volunteer investigations showed that anatomical site selection for wearable nanovibration delivery must balance measurable transmission against repeatability and practicality of application. Although the distal tibia showed the highest mean transmitted amplitudes in the initial investigation, this site was highly variable and difficult to attach the device to consistently because of the complex malleolar geometry. By comparison, the distal femur and proximal tibia provided more repeatable transmission, and in the revised prototype, the distal femur showed the highest mean transmitted amplitude. The distal femur was therefore taken forward because it combined clinical relevance with the most practical and stable configuration for external device attachment. More broadly, these findings show that successful translation of a wearable mechanical intervention depends not only on delivered signal magnitude, but also on interface conformity, positioning reproducibility, and usability in the intended patient setting.

The bovine femur experiments provided direct evidence that externally applied nanovibration at 1 kHz can be transmitted across and into bone under controlled benchtop conditions. Surface mapping showed that the highest amplitudes occurred in the region directly opposite the transducer, while depth-resolved measurements confirmed that measurable nanoscale vibration remained detectable within the condylar region rather than being confined to the immediate surface. This suggests that the condylar region vibrated as a rigid body, rather than transmission occurring through other means (e.g. surface waves travelling only through the cortical bone layer). Although only a small fraction of the transducer displacement was transmitted into the bone, normalised measurements indicated that this corresponded to approximately 1–2 % of the transducer surface amplitude, and the retained signal remained within the nanoscale range at the target operating frequency.

This data also showed that interface conditions were a major determinant of transmitted vibration. The gel interface layer altered the measured signal within bone, indicating that the delivered bone-level vibration depends on the coupling conditions between the transducer and the tissue rather than simply on the transducer output. Because the gel interface layer was included in the wearable device to reduce pressure-related tissue risk, these findings highlight a key translational trade-off in the current design: improving safety and interface conformity can reduce the transmitted mechanical dose. These observations are consistent with the broader bone-conduction literature, which shows that externally applied vibration can propagate through osseous structures and that measured transmission is strongly influenced by anatomy, contact conditions, frequency, and intervening soft tissues (Stenfelt et al., 2000).

The reduction in vibration amplitude from the transducer to the bone is likely due to two factors. Firstly, the mass of the transducer itself will impact the transfer of motion to the leg. The mass of the transducer used was 226 grams, whilst the mass of a leg is typically >10 kg. This significant difference in inertial masses is therefore expected to be the primary limitation in amplitude transfer. Secondly, the preloading conditions of the transducer will provide additional counterforce to increase the vibration amplitude in the condylar region, with elastic strapping as the preload source. Greater preloading may increase vibration transmission to the bone, but this must be balanced against the actuator’s blocking force and optimal preload conditions.

The repeat-application measurement on bovine bone at 1 kHz further showed that transmitted vibration was sensitive not only to the presence or absence of the gel layer, but also to reapplication-related factors such as positioning, strap tension, local conformity, and possible device slippage. This is important translationally because a wearable intervention will inevitably be removed and reapplied across sessions, meaning that the effective bone-level dose may vary even when the nominal transducer settings are unchanged. In our testing, variability due to repeated application was minimised by using the gel interface layer.

The preliminary clinical feasibility evaluation showed that localised nanovibration could be delivered and monitored repeatedly in the intended SCI setting without evidence of pressure-related skin injury. The repeated-session dataset demonstrated that the intervention could be applied over an extended period while maintaining measurable nanoscale vibration at the distal femur during real-world use. These findings support the practical use of the device in the intended setting, but do not address therapeutic efficacy.

At the same time, the feasibility study highlighted important barriers to routine use with the current design. Session duration appeared to plateau below the intended maximum, suggesting that longer periods of continuous wear were not yet practical. Gradual device slippage, audible transducer noise, and the bulk of the external electronics and cabling all reduced convenience during use, indicating that further engineering development will be needed before the system becomes a more usable clinical intervention.

This study has several important limitations. First, the clinical feasibility evaluation was completed by only one participant, which precludes any inference regarding clinical efficacy or generalisability. Second, although the bovine femur experiments provided useful benchtop evidence of vibration transmission across and into bone, bovine bone is only a proxy for the *in vivo* human lower limb and does not reproduce the full anatomical, soft-tissue, and physiological environment. Third, the delivered bone-level signal was strongly influenced by interface conditions, indicating that the effective in vivo dose cannot be inferred directly from transducer output alone. Finally, some measured amplitudes approached the practical limits of resolution, and therefore, small numerical differences should be interpreted cautiously.

These limitations point to two priorities for future work. The first is further device optimisation to improve wearability and mechanical coupling in the intended clinical setting. The second is translational evaluation in a larger SCI cohort, with bone imaging outcomes incorporated to assess whether repeated localised nanovibration produces measurable skeletal effects. In parallel, the lower amplitudes achieved with the present wearable system may warrant additional *in vitro* validation, given that prior stem-cell studies focused on higher amplitudes. Together, these steps would enable the current proof-of-concept platform to progress toward a more rigorous clinical investigation.

In summary, this study shows that a wearable system can be used to deliver and monitor localised nanovibration at the distal femur, and that nanoscale vibration can be transmitted into bone under controlled conditions. Although therapeutic efficacy remains unproven, the work establishes a practical translational foundation for further development of localised wearable nanovibration as a mechanically targeted intervention for fracture-prone skeletal sites after SCI.

## Acknowledgements

The authors thank the participants and staff of the Queen Elizabeth National Spinal Injuries Unit for their support of the clinical feasibility evaluation.

## Author Contributions

J.A.W, R.G., P.C., M.P., S.C., and S.R. conceived the experiments. R.G. and J.A.W. performed the experiments. J.A.W and R.G. analysed the data. R.G. designed the wearable harness. P.C. designed the electronics. P.G.C., S.C., J.S.R., M.P., M.J.D., and S.R. contributed expertise and supervision. M.P., S.C., J.S.R. and S.R. acquired funding. J.A.W. wrote the manuscript and prepared the figures. All authors have given approval to the final version of the manuscript.

## Data Availability Statement

The data that support the findings of this study are available from the corresponding author, S.R, upon reasonable request.

## Ethical Standards

The authors assert that all procedures contributing to this work comply with the ethical standards of the relevant national and institutional committees on human experimentation and with the Helsinki Declaration of 1975, as revised in 2008. Ethical approval for the human study was obtained from NHS Greater Glasgow and Clyde / West of Scotland REC 3 (REC reference 15/WS/0009; IRAS project ID 84046), and all participants provided informed consent.

