## Supplementary Material for "Development and Preliminary Clinical Feasibility of a Wearable Nanovibration Delivery Device for Localised Bone Stimulation in Individuals with Spinal Cord Injury"

**Contents**

**Supplementary Material A - Patient device electronic system description**

**Supplementary Material B - Device configuration for the second healthy volunteer nanovibration measurements**

**Supplementary Material C – Bovine femur Depth-resolved vibration transmission experimental setup**

**Supplementary Material D - Device instruction manual**

**Supplementary Material E - Depth-resolved transmission normalised to transducer output.**

**Supplementary Material A - Patient device electronic system description**

The following sections describe the electronic systems implemented for the patient nanovibration delivery device, including generation of the 1 kHz drive signal for the bone conduction transducer, amplification of the ACH-01 accelerometer output, portable processing and logging of the measured signal, power regulation, and accelerometer calibration.

The patient device was designed as a portable, stand-alone system that could be issued to a participant for home or ward-based use with minimal user interaction. The system comprises a wave generation and drive stage for the bone conduction transducer, an amplification and conditioning stage for the ACH-01 accelerometer, a microcontroller-based data logging and user-feedback module, and a battery-powered voltage regulation stage.

The wave generator circuit is based on the one used to generate a sine-wave voltage signal in the Nanokick bioreactor (Campsie et al., 2019). An AD9833 low-power, programmable waveform generator (Analog Devices, Massachusetts, USA) produces the required sine wave with the output frequency and phase programmed using an ATMega328 microcontroller (Atmel, California, USA). Filtering is required at the output of the AD9833 to significantly reduce higher-frequency components generated during the digital synthesis of the primary signal, using a seventh-order LC elliptical reconstruction filter. A non-inverting amplifier circuit, using an OPA37 ultra-low noise OP-AMP (Texas Instruments, Texas, USA), is utilised to boost the amplitude of the filtered sine wave before it reaches the final amplification stage. A 10 kΩ rotary potentiometer in the non-inverting amplifier circuit allows the gain to be adjusted by the user. Finally, the sine wave signal is amplified with a MAX98306 3.7W power amplifier (Maxim Integrated, California, USA) to provide the bone conduction transducer with the voltage and current required to function. The power amplification stage was purchased as a standalone printed circuit board (PCB) (Adafruit Industries, New York, USA). A block diagram of the circuitry described above is shown in Figure A1, and the wave generator and power amplifier PCBs are shown in Figure A2.

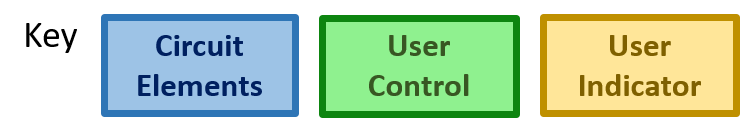

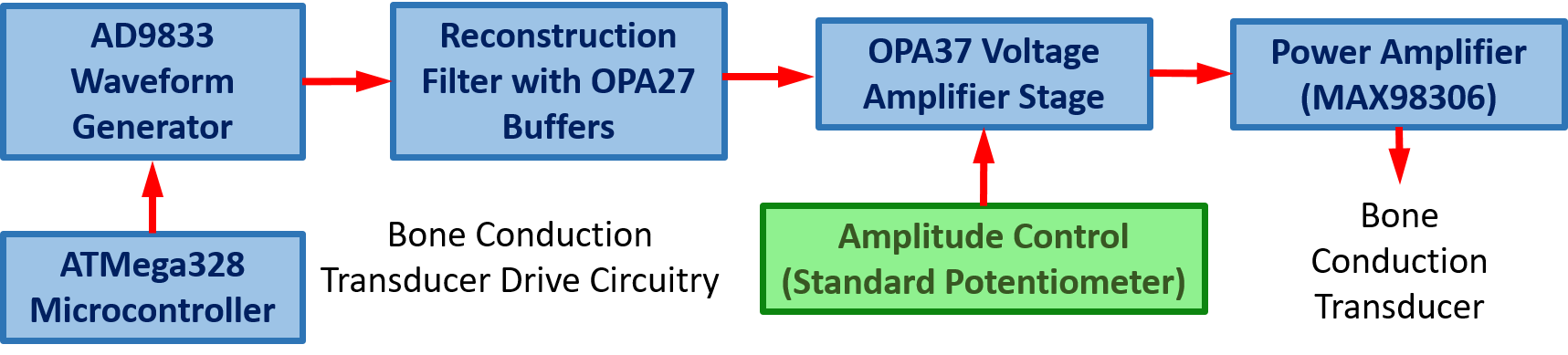

**Figure A1**: Block diagram of main components of the wave generator PCB used in the animal study.

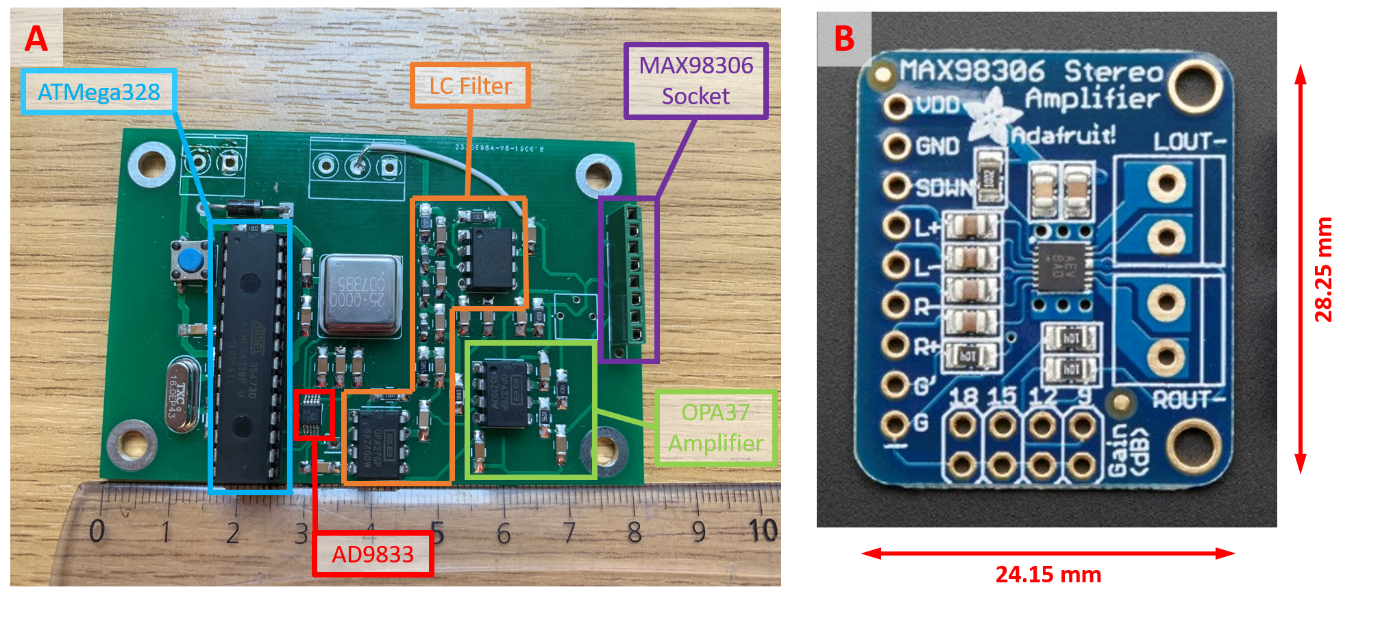

**Figure A2**: (A) Photograph of the wave generator PCB with main components highlighted (B) Image of power amplifier (MAX98306) PCB purchased from Adafruit Industries.

The vibration transmitted through the target anatomical region is detected using an ACH-01 accelerometer (TE Connectivity, Schaffhausen, Switzerland). Because the accelerations being measured are extremely small, the ACH-01 output is first amplified using the OPA37-based multi-stage amplifier circuit. Rather than sending this signal to a PC-based acquisition system, as in earlier laboratory development work, the amplified sine wave is converted to a DC signal using an AD736JNZ RMS-to-DC converter (Analog Devices, Massachusetts, USA) before being processed by an Arduino Nano microcontroller (Arduino, Massachusetts, USA). The Arduino applies the appropriate calibration equation to the measured DC voltage to convert it into displacement and saves the displacement data to a text file on a micro SD card. The micro SD card interface was purchased as a standalone PCB (Adafruit Industries, New York, USA). The Arduino also assesses whether the detected displacement is within the acceptable operating range and illuminates a red LED when it is outside this range (< 1.5 nm or > 40 nm) and a green LED when it is within range, allowing the user to confirm correct operation and, where required, adjust the transducer output using the rotary potentiometer in the drive circuit. A block diagram if this system is shown in Figure A3.

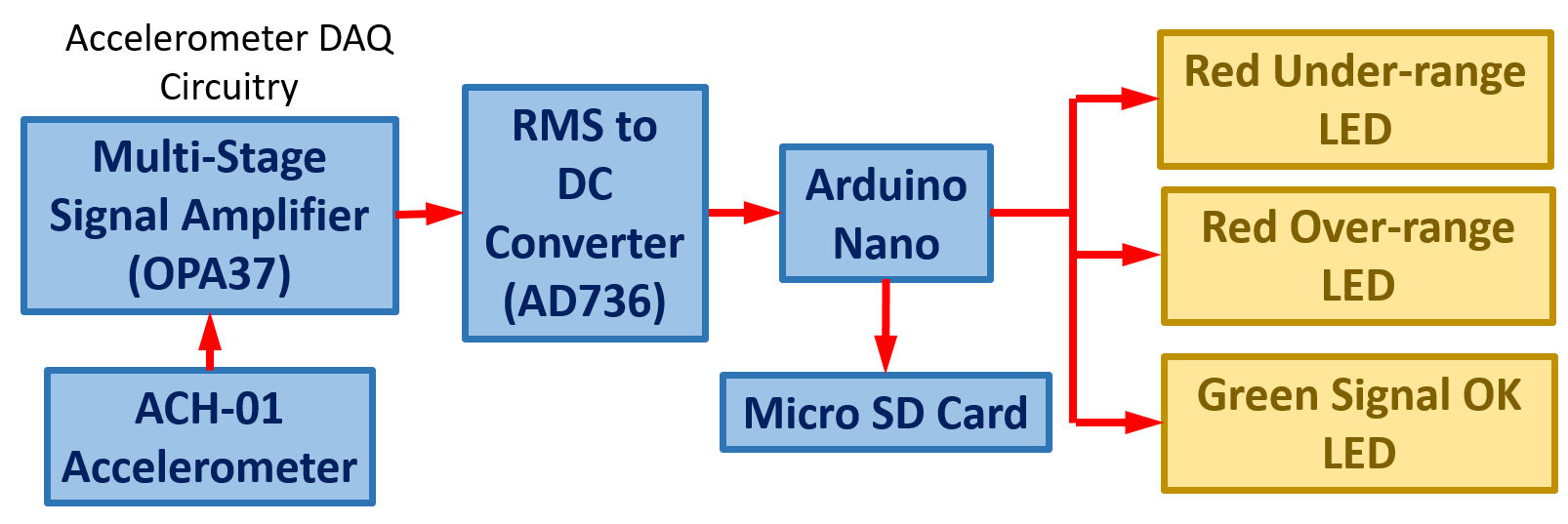

**Figure A3:** Block diagram of the main components of the accelerometer DAQ system used in the patient study.

The patient device needs to be portable; therefore, the circuitry needs to be battery-powered and easily rechargeable. A portable phone charger (Jonkuu, Shenzen, China), that outputs a constant 5V DC with a charge of 10000mAh, was used as the primary power source and the voltage output from this source was adjusted to the appropriate levels required throughout the system using boost converters, DC-DC converters or linear regulators. A block diagram of the voltage regulation is shown in Figure A4.

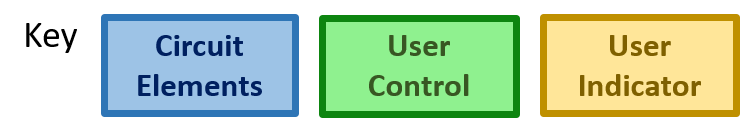

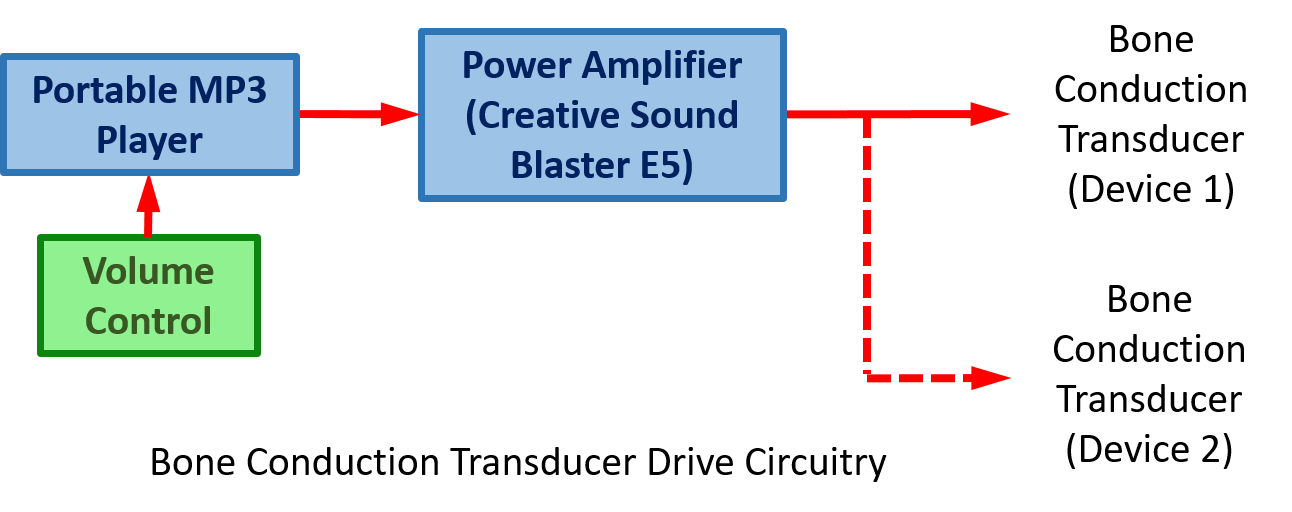

**Figure A4**: Block diagram of a more compact alternative system for the patient study. The system is in the early stages of testing and has not been used on a participant yet.

**Calibration of ACH-01 accelerometer**

Each ACH-01 accelerometer produces a slightly different voltage output for a given acceleration because of device-to-device variation arising from the manufacturing process. Therefore, all accelerometers used with the patient device were individually calibrated and assigned their own calibration curve. It was also important to express the output in terms of displacement rather than acceleration so that measurements were directly comparable with previous nanovibration literature. The calibration process is carried out by magnetically fixing the accelerometer to a Nanokick bioreactor (Campsie et al., 2019), which creates a precise nanoscale oscillation at 1 kHz and different set amplitudes, and measures the displacement of the accelerometer with a laser interferometer (Model SP-SS IOS Meßtechnik GmbH, Ilmenau, Germany). The Nanokick bioreactor outputs a very precise and stable nanoscale vibration, using an array of piezoelectric ceramics, for cell culture experiments and is itself calibrated by laser interferometry, an instrument that can measure displacements with sub-nanoscale resolution. Before calibration, each accelerometer is individually numbered using a scribe on the plastic casing so it is easily identified, a small section of self-adhesive rubber magnet is fixed to the underside of the accelerometer to attach to the bioreactor top plate, and retroreflective tape is attached to the top surface of the accelerometer to reflect as much of the interferometer’s laser light back to its’ sensor as possible for an accurate measurement. The bioreactor is driven by a 1 kHz sine-wave signal generated by an AFG-21005 arbitrary function generator (GW Instek, New Taipei City, Taiwan) and amplified by a Behringer KM750 amplifier (Behringer, Willich, Germany). The frequency is fixed at 1 kHz, and the displacement amplitude is adjusted by changing the output voltage amplitude on the function generator. Two sets of calibration measurements were taken because it was found that over a measurement range of 1 – 75 nm the calibration curve does not remain exactly linear. For wide range measurements, the amplitude of the sine wave is incrementally increased from 2.5 nm to 75 nm, and the ACH-01 voltage is measured with an oscilloscope (Vrms and Vpk-pk), see Figure A5B, and for shorter range measurements, data is taken from 1 – 10 nm in incremental steps of 1 nm, see Figure A5A. It was determined that the calibration curve for the lower-range measurements would yield the most accurate results. This is the exact same calibration process used in a previous study, were we developed a wearable nanovibration device for a rat model of complete SCI (Williams et al., 2024).

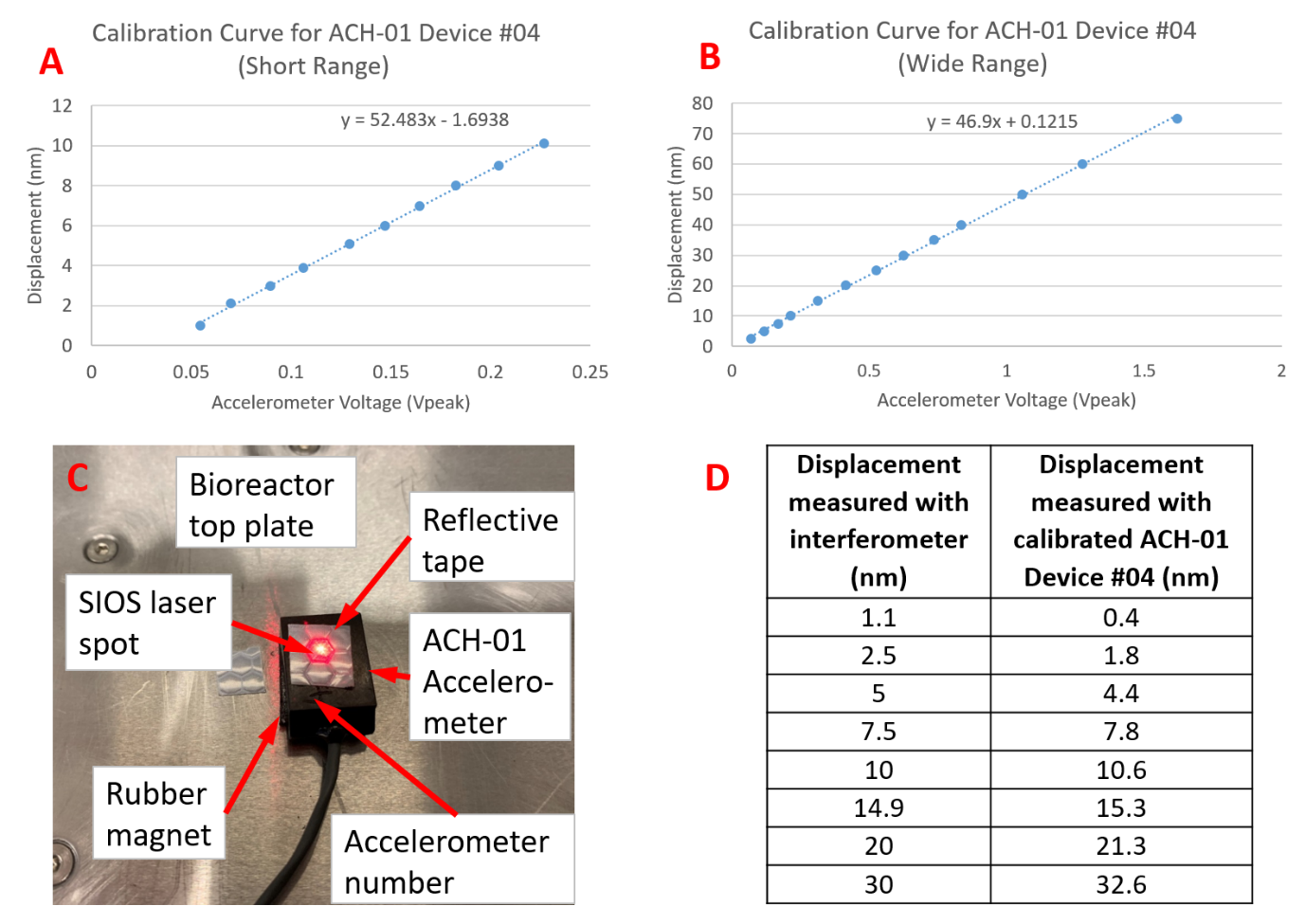

**Figure A5**: (A) Plot of the calibration data for ACH-01 device 4 over a short range of displacement values (B) Plot of the calibration data for ACH-01 device 4 at the higher range of displacement values (C) Picture of ACH-01 device 4 on bioreactor top plate being measured with the laser interferometer (D) Comparison of displacement measured with interferometer with data recorded by Spike2 software from calibrated ACH-01 device 4.

**Supplementary Material B - Device configuration for the second healthy volunteer nanovibration measurements**

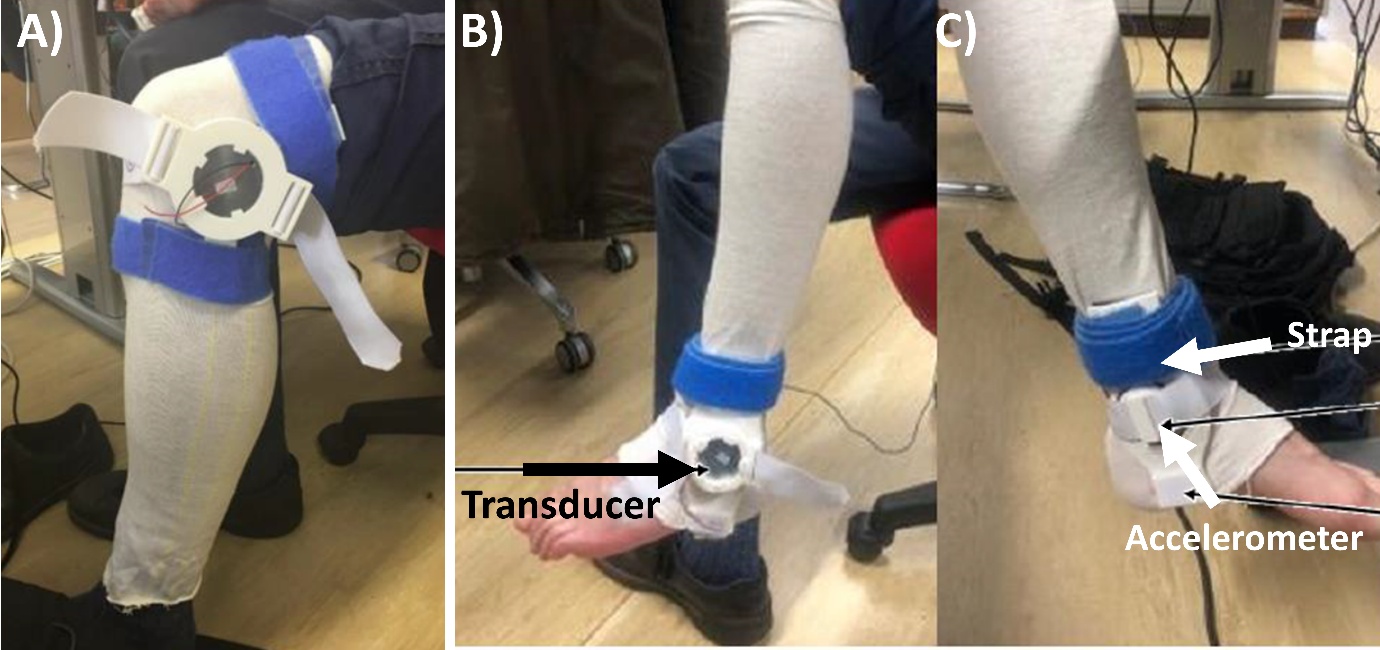

**Figure B1. Device configuration for the second healthy volunteer nanovibration measurements.** (A) Photograph of the wearable nanovibration device applied at the distal femur and proximal tibia during surface transmission measurements. (B–C) Photographs of the ankle version positioned at the distal tibia (ankle), showing lateral (B) and medial (C) views. The bone-conduction transducer was positioned on the lateral aspect of the limb, with the accelerometer on the medial side, and the device secured using elastic strapping.

**Supplementary Material C – Bovine femur depth-resolved vibration transmission experimental setup**

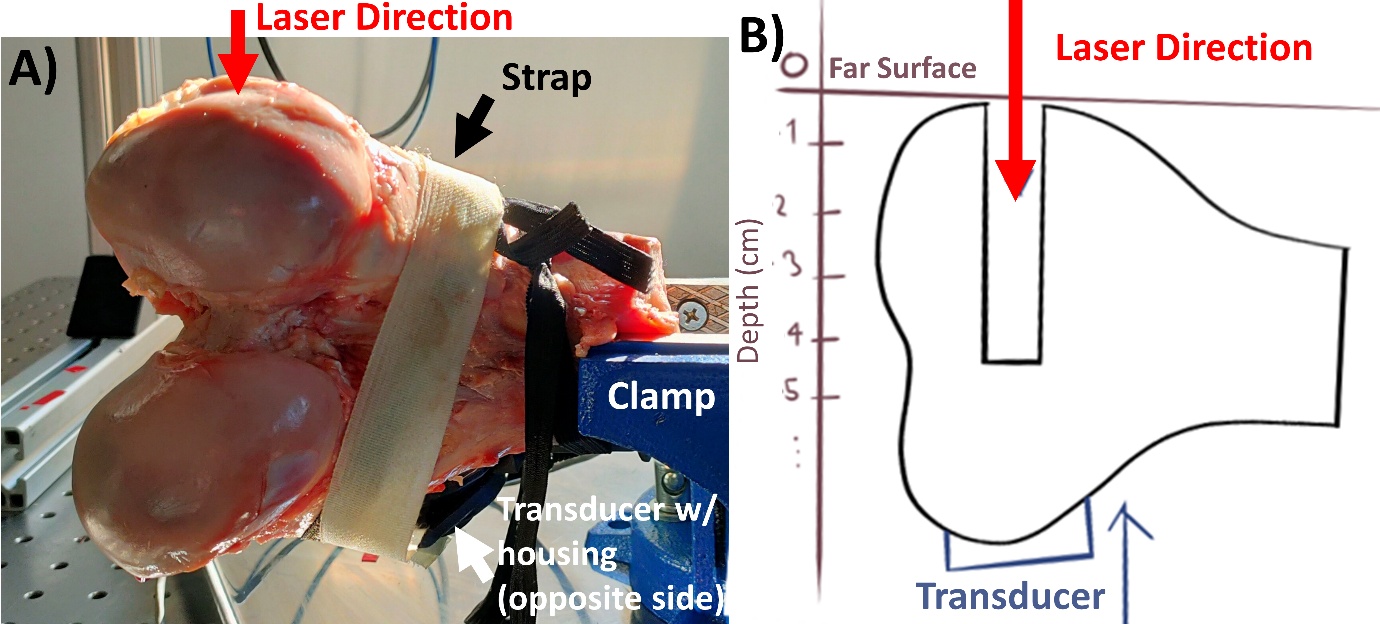

**Figure C1. Experimental setup for depth-resolved vibration transmission in bovine femur.** (A) Photograph of the experimental arrangement showing a bovine distal femur secured in a clamp, with the nanovibration voicecoil transducer applied to the condylar surface using elastic strapping and a compliant interface layer. (B) Schematic representation of the measurement configuration. A channel was drilled through the condylar region in 1 cm increments, and reflective tape was placed at each depth to enable measurement of vibration amplitude using a single-point laser vibrometer. Vibration was applied laterally via the transducer, and measurements were taken along the drilled path to assess depth-resolved transmission through bone.

**Supplementary Material D – Device instruction manual**

**Instructions for Knee Nanovibration Device**

**Contact Information:**

**Email:	Telephone:**

**Note: These instructions are for the right leg**

**Step 1:** Open straps up and lay out as shown.

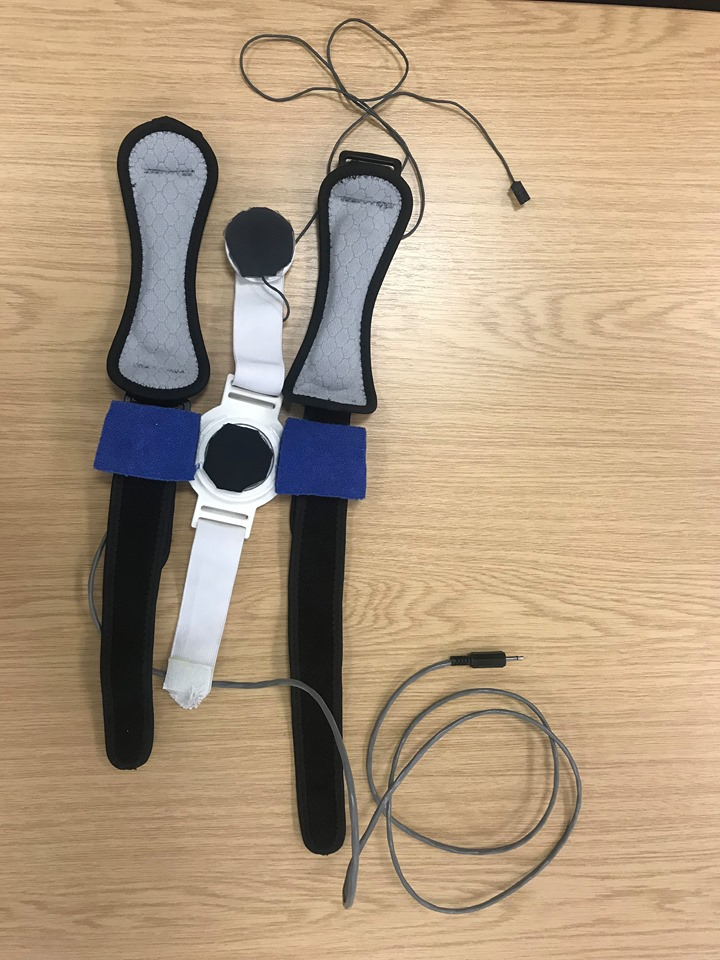

**Wire A**

**Black Padded Strap**

**Elasticated White Strap**

**Central Hub (C)**

**Strap Arms**

**Wire B**

**Step 2:** Adjust device against the outer leg as shown.

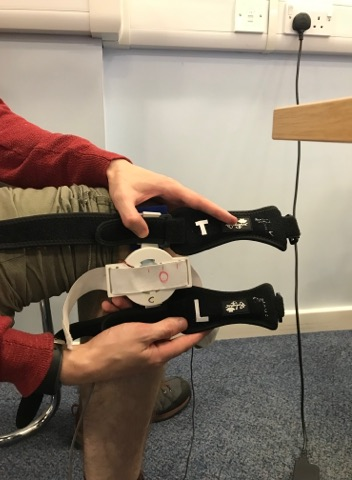

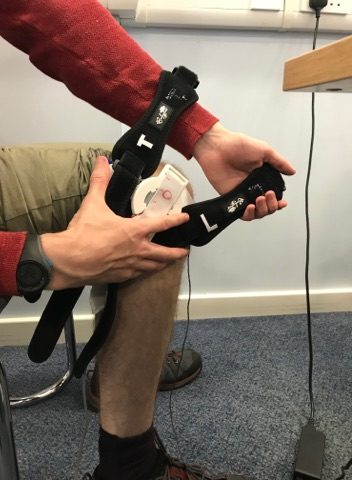

**Step 3:** Position the top strap (labelled T) on the top of the thigh as shown below. The central hub should be positioned against the outer leg and the padded part of the strap should be above the knee. Close the strap at the inside of the leg and tighten it lightly so as to secure the device from falling.

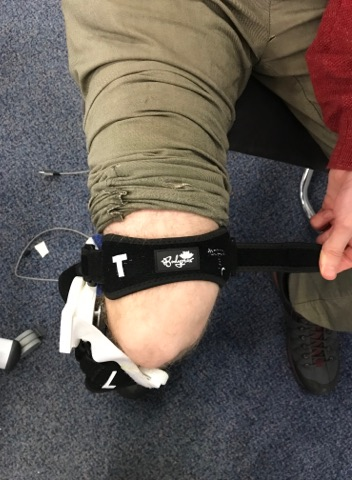

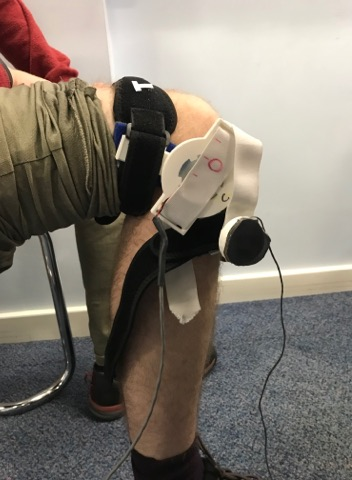

**Step 4:** Position lower strap (labelled L) on the lower leg below the knee. As with in step 3, close the strap at the inside of the leg and tighten lightly.

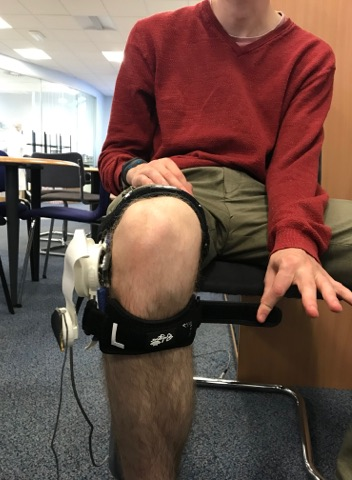

**Step 5:** Rotate central hub so that the white strap passes evenly in-between the two black straps as shown.

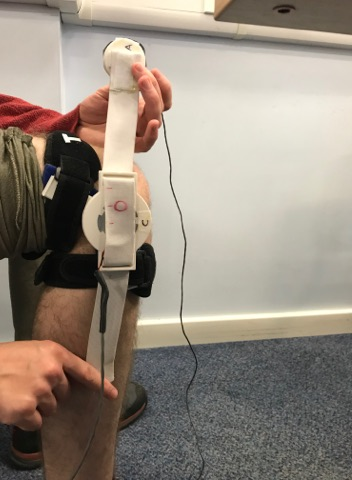

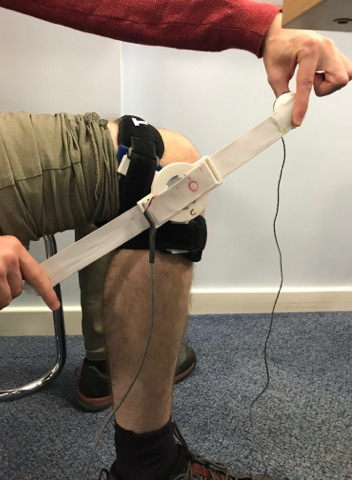

**Step 6:** Shift central hub to be positioned at the end of the lower leg above the knee joint.

**Step 7:** Position the part A at the end of the white strap against the knee on the opposite side of the central hub. Attach the other end of the white strap on top of the part A to secure in place.

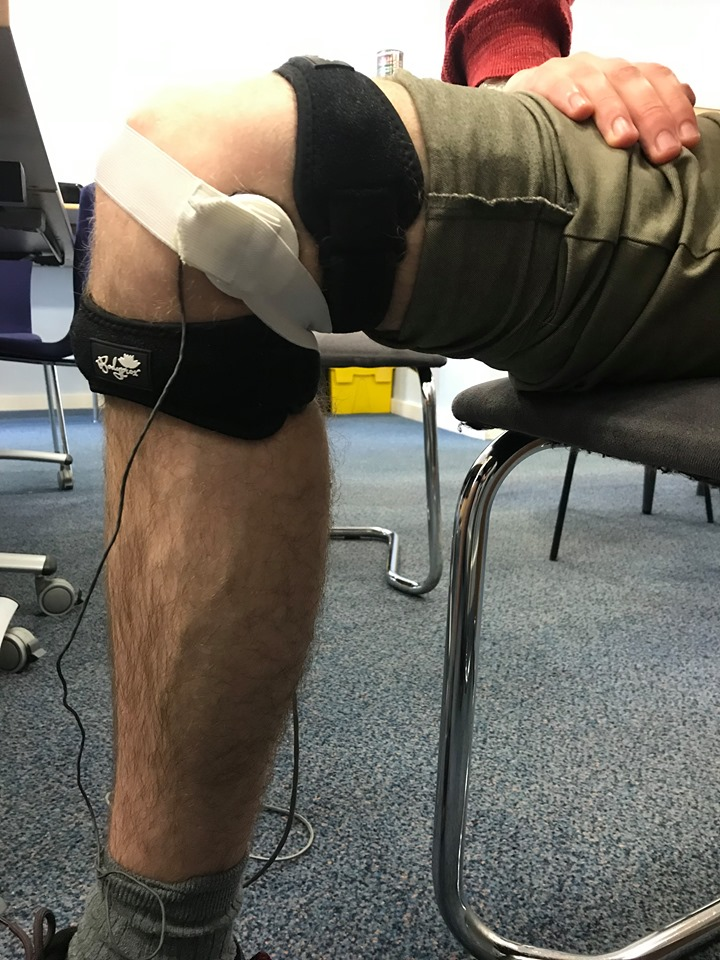

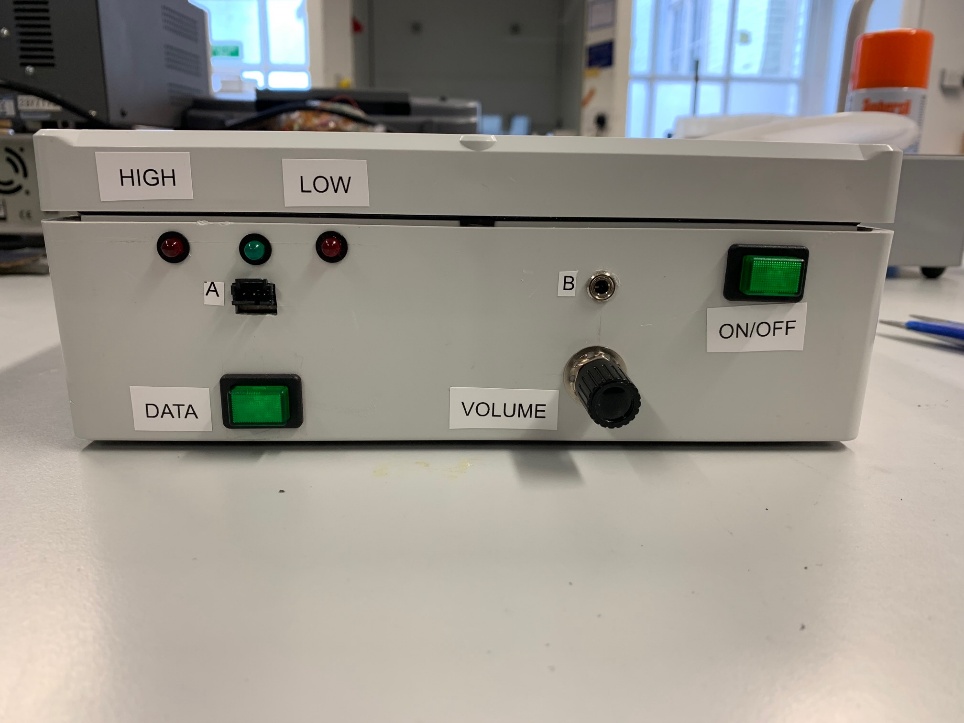

**Step 8:** Connect wires A and B as shown below

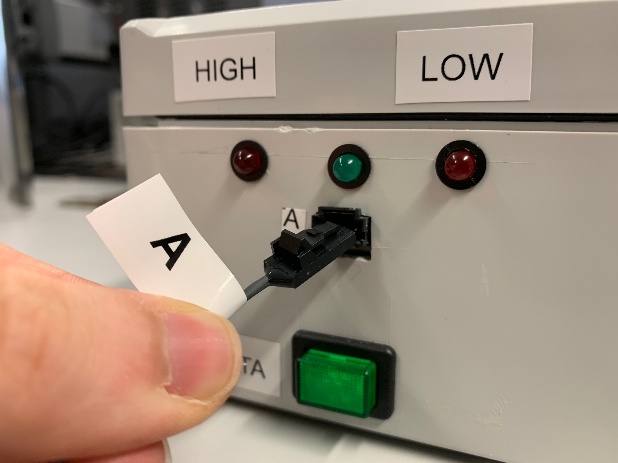

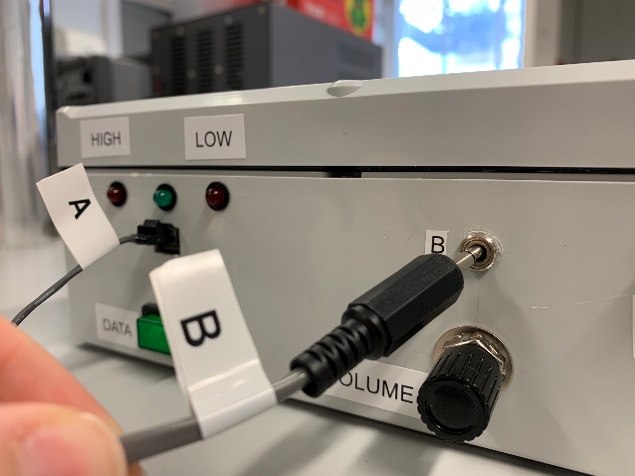

**Step 9:** Turn on the power supply using the ON/OFF button. The red and green lights on the left side of the device will flash for a few seconds to show that the power is on.

If both red lights continually flash, the unit should be turned off and the issue reported to one of the contacts above.

**Step 10:** If the green light is on then the setup is good and you can move to step 11.

If the red light on the left is on, then the signal is too high. The signal can be turned down by turning the VOLUME control to the left. If the left red light is still on, loosen straps slightly until green light comes on.

If the red light on the right is on, then the signal is too low. The signal can be increased by turning the VOLUME control to the right. If the red light is still on after turning the volume control all the way to the right, then the device on the leg needs to be readjusted (see step 6)

**Step 11:** Once the setup is complete, press the DATA button on the left side of the power supply to start recording data.

**Step 12:** After the time for the therapy is complete press the DATA button to stop recording data.

**Step 13:** Press the ON/OFF button to power down the device and disconnect the wires A and B.

**Step 13:** Remove the white strap by detaching part A.

**Step 14:** Remove the lower black strap.

**Step 15:** Remove the upper black strap slowly to ensure that the device does not fall, pull device away from leg.

**Step 16:** Plug in USB cable as shown below and plug to mains. After a 4-hour session, make sure to charge the device for at least 4 hours.

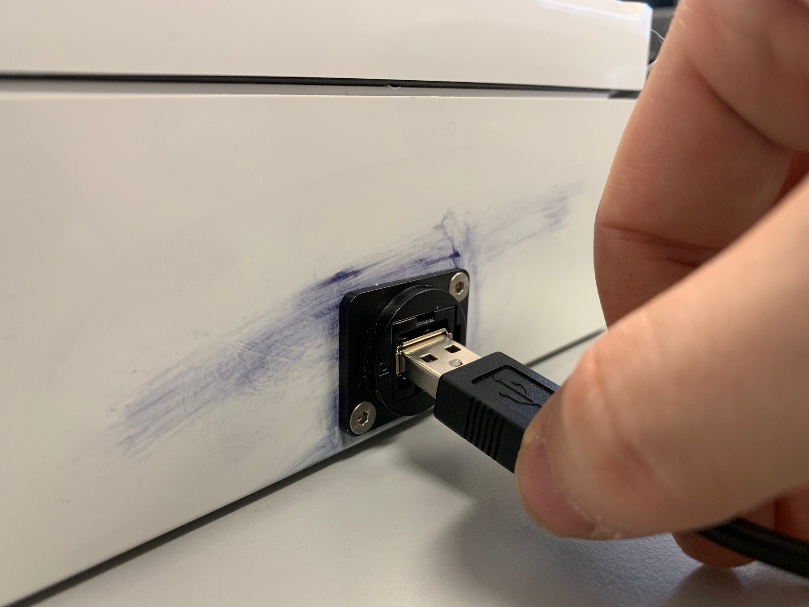

**Supplementary Material E - Depth-resolved transmission normalised to transducer output.**

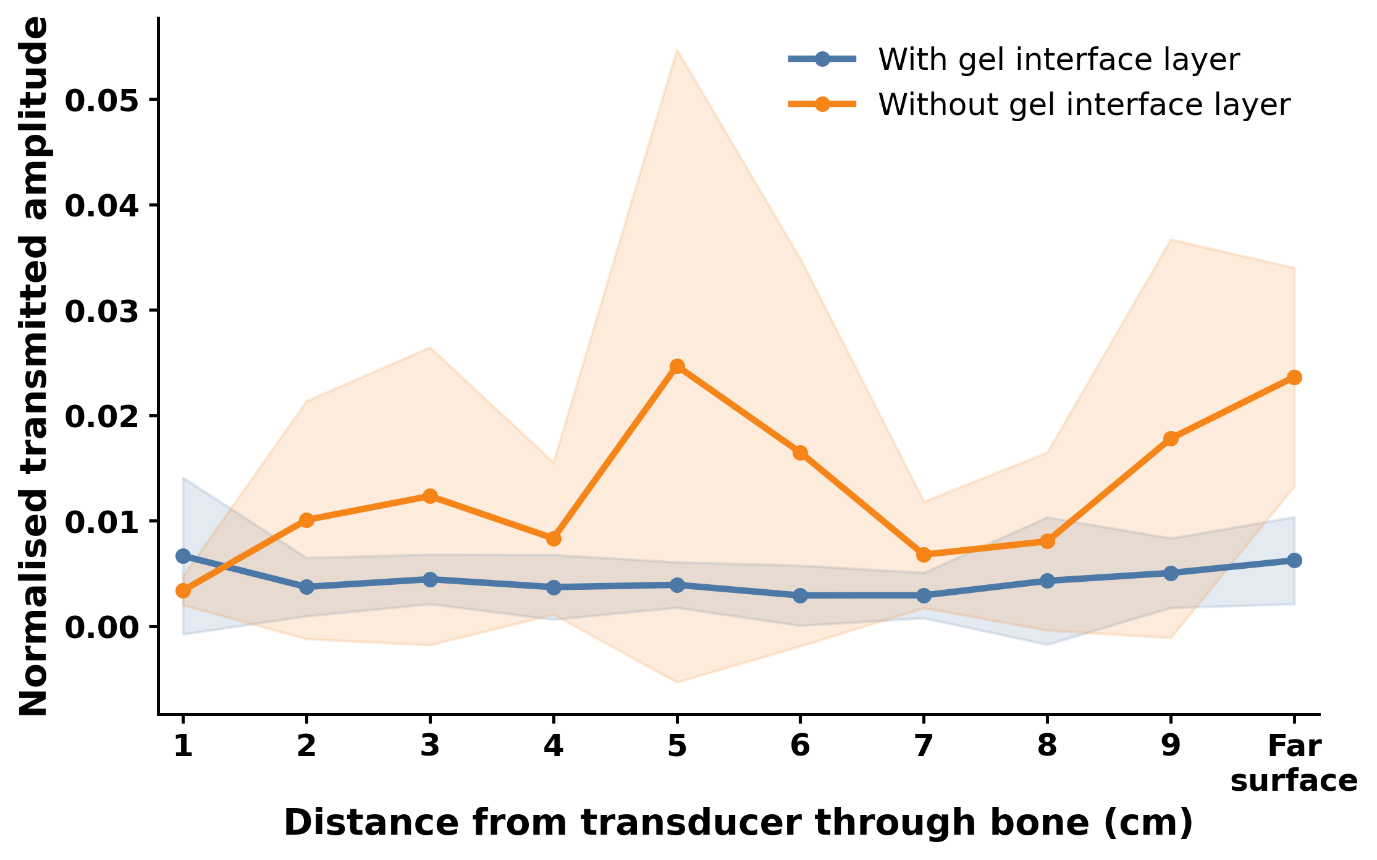

**Figure E1. Depth-resolved transmission normalised to transducer output.** Mean transmitted vibration amplitude (± SD), normalised to the corresponding transducer surface amplitude measured under direct coupling to the bone, is shown as a function of distance from the transducer. Depth is defined as the distance through bone from the transducer-side surface (0 cm) to the far cortical bone surface.
